# Determinants of Completeness and Timeliness of Pneumococcal Conjugate Vaccine Immunization in Yogyakarta, Indonesia: A Cross-Sectional Study

**DOI:** 10.64898/2026.02.03.26345526

**Authors:** Noorkhalisah Noorkhalisah, Risalia Reni Arisanti, Solikhin Dwi Ramtana, Mei Neni Sitaresmi

## Abstract

Pneumonia remains a leading cause of global child mortality. Following the Pneumococcal Conjugate Vaccine (PCV) introduction in Yogyakarta, Indonesia, uptake for the primary series (PCV1 and PCV2) exceeded 90%. However, PCV3 coverage remained suboptimal (60% in 2023; 75% in 2024), indicating significant dropout. This study aimed to identify determinants of PCV immunization completeness and timeliness to address this gap.

We conducted a cross-sectional study using cluster sampling among 405 caregivers of children aged 13–37 months in Yogyakarta City in March 2025. Data were collected via structured digital questionnaires assessing socio-demographics, perinatal conditions, knowledge, support systems, and attitudes toward multiple injections. Multivariate logistic regression was employed to determine factors associated with PCV immunization completeness and timeliness.

Of 398 participants (98.3% response rate), the majority were female (95.7%) and housewives (75.1%). The prevalence of PCV completeness was 66.3%, while timeliness was only 36.4%. Multivariate analysis revealed that acceptance of multiple injections was the strongest predictor for both completeness (aOR 49.18; 95% CI: 21.30–113.50) and timeliness (aOR 22.04; 95% CI: 6.55–74.08). Additionally, home ownership (aOR 1.93; 95% CI: 1.04–3.58) was associated with completeness, whereas high knowledge (aOR 1.85; 95% CI: 1.12–3.03) improved timeliness. Conversely, preterm birth was significantly associated with lower odds of timeliness (aOR 0.29; 95% CI: 0.09–0.88).

Acceptance of multiple injections emerged as the most critical modifiable factor for both outcomes. To optimize the PCV program, health authorities should prioritize counselling strategies to alleviate parental concerns regarding multiple injections. Additionally, intensified monitoring for preterm infants is crucial to mitigate immunization delays.

## INTRODUCTION

Pneumonia is the single largest cause of death from infection in children worldwide. Globally, there are around 1,400 cases of pneumonia per 100,000 children, or one case per 71 children annually [1]. Children under five years of age, particularly those under two, are at greatest risk of infection and death [2]. In 2019, pneumonia accounted for 740,180 (14%) of deaths among children under 5 years of age, or approximately 2,200 died every day, 92 children per hour, and 1 child every 39 seconds due to pneumonia [3–5].

*Streptococcus pneumoniae* is the predominant pathogen in paediatric community-acquired pneumonia and the leading cause of global pneumonia mortality, accounting for 18% of severe cases and 33% of pneumonia-related deaths worldwide [6]. Mortality and disease burden are disproportionately higher in developing countries, particularly in South Asia and sub-Saharan Africa[3]. In Indonesia, the prevalence of pneumonia among infants is 4.8%, peaking at 6% among those aged 12–23 months [7], with an estimated incidence of 3.55% [8]. Pneumonia is the second leading cause of death overall and the primary cause of mortality among Indonesian children under five, contributing to 15.3% of deaths [9]. With nearly 20,000 childhood pneumonia deaths annually, Indonesia has the highest burden in Southeast Asia[4].

Pneumococcal Conjugate Vaccines (PCV) were introduced into the infant immunization program to reduce the burden of pneumococcal disease in children. PCV immunization significantly reduced the prevalence of invasive pneumococcal disease in children[6]. The first efficacy trial in Northern California showed nearly 100% vaccine effectiveness against invasive pneumococcal infections in children [10]. Although pneumonia remains the leading cause of post-neonatal mortality in Indonesia, studies have confirmed that introducing PCV immunization would be a highly cost-effective intervention for the national program [11].

The World Health Organization (WHO) recommends the inclusion of the Pneumococcal Conjugate Vaccine (PCV) into National Immunization Programs (NIP), particularly in countries with high under-five mortality. Indonesia began phased PCV introduction in 2020, with national implementation in 2022. During the 2017–2019 demonstration phase, coverage averaged above 80%. The program employs the 13-valent PCV, administered at 2, 3, and 12 months of age, effectively reducing pneumococcal serotypes responsible for severe pneumonia [12].

PCV immunization in Yogyakarta City commenced on September 13, 2022, targeting infants born on or after June 13, 2022. The first PCV1, at 2 months) and the second (PCV2 at 3 months) doses are co-administered with other immunizations, including the DPT-HB-Hib vaccine, oral polio vaccine (bOPV), oral rotavirus vaccine, and IPV vaccine (specific to the Special Region of Yogyakarta). The booster dose (PCV3) is administered at 12 months of age [13]. Despite high coverage for PCV1 (95.93%) and PCV2 (94.14%), PCV3 coverage in 2023 was markedly lower at 59.87%. This gap is concerning, as PCV3—part of the national booster immunization schedule—is not co-administered with other vaccines, which may contribute to reduced uptake. Therefore, this study aims to analyse the factors influencing the completeness and timeliness of Pneumococcal Conjugate Vaccine (PCV) immunization in Yogyakarta City in 2024.

## METHODS

### Study Design, Setting, and Sampling

A quantitative descriptive study with a cross-sectional design was conducted in Yogyakarta City from March 6 to 27, 2025. The study population included children aged 13–37 months from the 2022–2023 birth cohort recorded in the local immunization database. The minimum sample size was determined using OpenEpi [14], assuming a 95% confidence level, 5% margin of error, design effect of 1.5, and a 10% non-response rate. Based on a total eligible population of 2,568 children for Pneumococcal Conjugate Vaccine (PCV) immunization in 2022, a final sample of 405 parents or caregivers was obtained. Cluster sampling was applied, using neighbourhood units (*Rukun Warga*, RW) as clusters. Of 616 RWs across 45 sub-districts, 58 were randomly selected using the Research Randomizer tool. From each cluster, 5–9 respondents were recruited, yielding 405 participants. Data were collected through structured, validated questionnaires administered via face-to-face interviews by trained enumerators under the supervision of the principal investigator to ensure consistency and minimize interviewer bias.

### Eligibility Criteria

Children aged 13–37 months registered in the 2022–2023 immunization target cohort in Yogyakarta City were eligible for inclusion. This age range was selected to ensure that participants had the opportunity to complete the full pneumococcal conjugate vaccine (PCV) series (PCV1–PCV3), enabling assessment of both immunization completeness and timeliness. Eligible participants were those residing in the selected clusters (*Rukun Warga*, RW) during the study period whose caregivers provided informed consent. Children who had permanently moved out of the study area or whose caregivers were unavailable or declined participation were excluded.

### Study Variables

The dependent variables were immunization completeness - defined as receipt of all three PCV doses (PCV1–PCV3)—and timeliness, defined as vaccine administration at the recommended ages of 2 months for PCV1 [15], 3 months for PCV2 [15], and 12–15 months for PCV3 [16]. Independent variables included sociodemographic characteristics, perinatal factors of the child (gestational age, birth weight, mode of delivery, and duration of hospitalization), parental or caregiver knowledge and attitudes, family support, support from health workers, and attitudes and acceptance of multiple immunizations.

### Data Collection Tools

Primary data were collected using a structured questionnaire comprising closed-ended multiple-choice questions and checklists. The questionnaire was adapted from validated instruments used in previous studies and pre-tested for validity and reliability. Secondary data were obtained from maternal and child health record books and the Sistem Informasi Imunisasi Daerah Istimewa Yogyakarta (SIMUNDU) to verify each child’s immunization history, including multiple injections.

### Data analysis

Data were entered and cleaned in Microsoft Excel 365, then analysed in STATA version 17. The minimal anonymized data set underlying the findings of this study is provided in the Supporting Information (S1 Dataset). Univariate analysis described frequency distributions and percentages. Bivariate analysis was performed using the chi-square test or Fisher’s exact test when assumptions were not met. Multivariate analysis employed log-binomial regression, including variables with p<0.25 from the bivariate stage.

### Ethical Clearance

Ethical approval was obtained from the Research Ethics Committee, Faculty of Medicine, Public Health, and Nursing, Universitas Gadjah Mada (KE/FK/0182/EC/2025). Written and verbally informed consent was obtained from all participants, and the study complied with national and international ethical guidelines. Additional information regarding the ethical, cultural, and scientific considerations specific to inclusivity in global research is included in the Supporting Information (S1 Questionnaire).

## RESULTS

A total of 398 parents or caregivers were successfully interviewed, yielding a response rate of 98.3%.

### Sociodemographic and Child Characteristics of Respondents

Most parents of caregivers were female (95.73%), aged 30–39 years (42.36%), and unemployed (75.13%) (Table 1). Most had more than one child (65.32%) and lived in their own houses (71.61%). Most children were aged 18–23 months (56.53%), with a balanced sex distribution (51.26% male, 48.74% female). Most were born at term (90.95%) with normal birth weight (90.20%). Vaginal delivery was the most common mode of birth (57.04%), and most children were discharged within 0–7 days (97.24%).

**Table 1.**
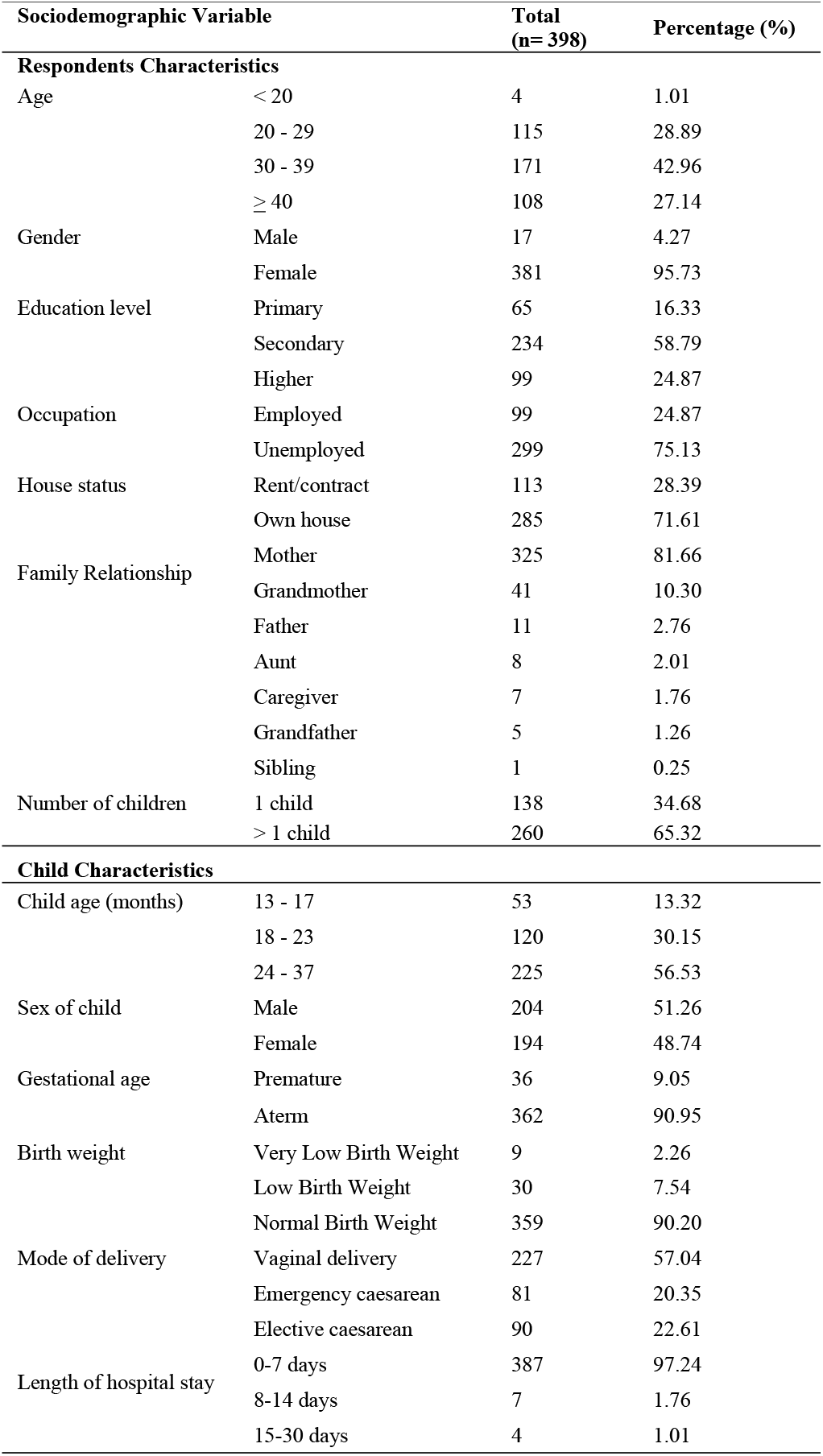
Showing Socio-demographic description of Respondents and Children.

### Completeness, Timeliness, and Acceptance of PCV Immunization among Respondents

**S1 Table**. Completeness, Timeliness, and Multiple Injection of PCV Immunization among Respondents

Completeness rates for PCV immunization were 83.17% (331 children) for PCV1 and 81.91% (326 children) for PCV2 (S1 Table). The overall series completeness (PCV3) was 67.09% (267 children), corresponding to a dropout rate of 16.08% from PCV1 to PCV3. Timeliness remained suboptimal, with 64.32% (256 children) receiving PCV1 on schedule and 56.03% (223 children) receiving PCV2 and PCV3 on time. Acceptance of multiple injections was high, with 77.64% (309 children) of caregivers consenting, while 22.36% (89 children) declined. Caregiver attitude toward multiple injections was a significant predictor of acceptance (OR = 2.05; 95% CI: 1.22–3.43).

### Barriers to PCV Immunization Completeness and Timeliness among Respondents

**S1 Fig. Barriers to PCV Immunization Completeness and Timeliness among Respondents** Barriers to immunization: An analysis of reported barriers revealed distinct patterns among groups. Lack of awareness or information was a prominent reason for incomplete immunization (33.67%), whereas it accounted for only 17.36% of cases in the delayed group. In contrast, the leading cause for delayed immunization was child illness (48.76%) (S1 Fig). These findings suggest that addressing information gaps is crucial for preventing dropouts, while health monitoring is key for improving timeliness.

### Bivariate Analysis

#### Factors Associated with Completeness and Timeliness of PCV Immunization

**S2 Table**. Bivariate Analysis of Factors Associated with Completeness and Timeliness of PCV Immunization

Bivariate analysis identified several factors associated with PCV immunization outcomes (S2 Table). Completeness was significantly associated with house ownership (OR=1.71; 95% CI: 1.05–2.74), elective caesarean delivery (OR=2.21; 95%CI: 1.22–4.11), parents or caregivers knowledge (OR=3.10; 95%CI: 1.95–4.91), attitudes of parents or caregivers (OR=1.52; 95%CI: 0.98–2.37), family support (OR=2.54; 95%CI: 1.40–4.59), and acceptance of multiple injections (OR=48.90; 95%CI: 21.68-122.32). Timeliness was associated with knowledge of parents or caregivers (OR=2.55; 95%CI: 1.65-3.99), family support (OR=2.13; 95%CI: 1.10-4.36), health worker support (OR=1.58; 95% CI: 0.98-2.54), and acceptance of multiple injections (OR=24.37; 95%CI: 7.75 – 122.44).

#### Multicollinearity assumption test

The analysis indicated no evidence of multicollinearity. All variance inflation factors (VIF) values are below the conservative threshold of 5, with the highest mean VIF recorded at 1.20.

#### Factors Associated with Completeness and Timeliness of PCV Immunization

Table 2. The results of the multivariate logistic regression show that several factors are associated with PCV immunization completeness and timeliness after full adjustment. Acceptance of multiple injections remained the strongest factor of completeness (aOR=49.18; 95% CI: 21.30–113.5), and timeliness (aOR = 22.04; 95% CI: 6.55–74.08). S4 Table. Living in an owned house was also significantly associated with completeness (aOR=1.92; 95% CI: 1.03–3.58). For timeliness, preterm birth (<37 weeks) was associated with a lower likelihood of timely immunization (aOR = 0.29; 95% CI: 0.09–0.88), and good caregiver knowledge increased the odds of timely vaccination (aOR = 1.85; 95% CI: 1.12–3.03).

**Table 2.** Multivariate Logistic Regression Analysis: Factors Associated with Completeness and Timeliness.

| Variable | Completeness | Timeliness |
| --- | --- | --- |
|  | aOR (95% CI) | aOR (95% CI) |
| **Age Respondents** |  |  |
| ≥ 40 | 1.84 (0.86 - 3.96) | 1.32 (0.74 – 2.33) |
| 20 - 29 | 0.97 (0.49 - 1.90) | 1.03 (0.58 – 1.82) |
| < 20 | 0.22 (0.02 - 1.78) | 1 |
| 30 - 39 |  |  |
| **Gestational age** |  |  |
| Premature | - | 4.55 (0.76 – 27.12) |
| Aterm |  |  |
| **House Status Respondents** |  |  |
| Own house | **1.92\* (1.03 - 3.58)** | 1.30 (0.75 – 2.23) |
| Rent/Contract |  |  |
| **Mode of delivery** |  |  |
| Elective caesarean | 1.74 (0.81 - 3.74) | 1.36 (0.77 – 2.38) |
| Emergency caesarean | 1.40 (0.65 - 2.99) | 1.23 (0.66 – 2.29) |
| Vaginal delivery |  |  |
| **Knowledge** |  |  |
| Good | 1.50 (0.82 - 2.72) | **1.85\* (1.12 – 3.03)** |
| Poor |  |  |
| **Attitude** |  |  |
| Positive | 1.20 (0.66 - 2.17) | **0.50\*\* (0.30 – 0.81)** |
| Negative |  |  |
| **Family Support** |  |  |
| Supportive | 1.32 (0.58 - 2.96) | 1.21 (0.57 – 2.54) |
| Less supportive |  |  |
| **Health worker support** |  |  |
| Supportive | 0.79 (0.42 - 1.49) | 1.14 (0.67 – 1.92) |
| Less supportive |  |  |
| **Attitude toward multiple injections** |  |  |
| Positive | 0.78 (0.39 - 1.52) | 1.34 (0.77 – 2.31) |
| Negative |  |  |
| **Acceptance of multiple injections** |  |  |
| Positive | **49.18\*\*\* (21.30 - 113.5)** | **22.04\*\*\* (6.55 – 74.08)** |
| Negative |  |  |

\*p < 0,05, \*\*p < 0,01, \*\*\*p < 0,001
Ref = Reference category

## DISCUSSION

This study demonstrated that two-thirds of children had completed the three-dose PCV schedule, yet only one-third received all doses on time. Multivariate analysis identified acceptance of multiple injections as the strongest factor consistently associated with both completeness and timeliness of PCV immunization. In addition, living in an owned house, which reflects higher SES, was positively associated with completeness, while good knowledge of parents or caregivers was associated with timeliness.

Preterm birth was negatively associated with timeliness, whereas family support and elective caesarean delivery showed associations in bivariate analysis but did not remain significant after adjustment. Acceptance of multiple injections emerged as the most dominant factor influencing both completeness and timeliness of PCV immunization. Bivariate and multivariate analyses consistently showed significant associations, indicating that parents who accepted multiple injections in a single visit were more likely to complete the PCV series on schedule. Clinical evidence further demonstrates that co-administration of DTaP-HepB-IPV with PCV is safe and well tolerated [17]. International recommendations also emphasize multiple injections, such as in Italy, where PCV is given simultaneously with hexavalent, rotavirus, and MenB vaccines to enhance coverage and adherence [18]. Studies in South Africa, The Gambia, and Albania showed that despite caregiver concerns, most children still received all recommended vaccines in a single visit [19–21]. Recent evidence further confirms that multiple injections are safe, effective, and broadly acceptable [1,22], underscoring their role in sustaining PCV and routine immunization coverage.

In terms of timeliness, findings from The Gambia showed that caregiver hesitancy rarely translated into delayed vaccination [20], while global guidelines emphasize that co-administration improves on-time coverage, particularly for children falling behind [23]. Furthermore, policy interventions in Ghana demonstrated that strengthening catch-up strategies and second year-of-life visits significantly improved timely vaccine uptake [24].

Parental knowledge was significantly associated with both completeness and timeliness of PCV immunization. Multivariate analysis confirmed that higher knowledge levels increased the likelihood of completing the PCV series and adhering to the recommended schedule. This finding supports prior evidence that adequate knowledge enhances vaccine uptake [25,26] and awareness of immunization benefits [27]. Studies also demonstrate that parental knowledge is strongly linked to complete immunization status[28] and full coverage in children aged 12–23 months[29]. Moreover, sufficient understanding of due dates has been shown to reduce vaccination delays, with knowledgeable caregivers having three times higher odds of timely vaccination [30].

Home ownership showed a significant association with PCV completeness but not with timeliness. In bivariate and multivariate analyses, caregivers living in their own homes were more likely to complete PCV immunization, although this effect was not consistent among parents. This finding aligns with previous evidence that stable residence facilitates proper maintenance of immunization records [31,32], while other studies reported no consistent association between home ownership and vaccination status [33]. In contrast, no significant effect was observed on timeliness, differing from evidence that urban residence is linked to on-time vaccination [30].

Family support showed a significant association with both completeness and timeliness of PCV immunization. Parents who received emotional and practical support were more likely to ensure full vaccination of their children. This aligns with previous studies reporting that family encouragement increases PCV uptake [25] and is significantly associated with complete primary immunization [28,34]. In addition, family support has been shown to correlate with timely vaccination, including BCG and other routine vaccines [35,36].

This finding is consistent with studies showing that delivery at health facilities increases the likelihood of timely vaccination [30,37] and that children born through caesarean section are more likely to receive neonatal vaccines completely [38]. However, other evidence suggests potential drawbacks, such as an increased risk of primary vaccine failure among caesarean-born infants [39].

The preterm infants often experience significant delays in initiating PCV, and other vaccines compared to term infants [40]. Nevertheless, safety studies confirm that PCV is well tolerated among preterm neonates, with immunogenicity comparable to that of term infants [41–43]. The findings underscore the importance of reinforcing provider confidence and ensuring adherence to recommended schedules, as prematurity should not be a reason to postpone immunization.

## CONCLUSIONS

This study identified key determinants of PCV immunization completeness and timeliness in Yogyakarta City. Acceptance of multiple injections and home ownership were significantly associated with PCV completeness, whereas parents’ or caregivers’ knowledge and acceptance of multiple injections also predicted timely vaccination. In contrast, preterm infants (<37 weeks) had lower odds of receiving PCV on schedule, highlighting persistent vulnerability in this group. These findings emphasize that strategies to strengthen parental awareness, family involvement, and acceptance of multiple injections are crucial to improving both coverage and timeliness.

Health authorities should strengthen Information, Education, and Communication (IEC) using engaging media, reminders, and clear guidance for pre-term infants. Primary health centers need to embed IEC into routine services, send booster reminders, and train staff to address concerns about multiple injections. Families are encouraged to support reminders and trust the safety of PCV. Future studies should use mixed methods to explore social and psychological determinants with direct decision-makers as respondents.

## Data Availability

Yes, data is available in Supporting Information: S1.Dataset

## Acknowledgements

We would like to thank the Yogyakarta City Health Office and the Provincial Health Office of the Special Region of Yogyakarta for their collaboration in this study. We are also grateful to the primary health center staff, immunization officers, enumerators, and community health cadres for their valuable assistance in collecting the data.

## Authors’ Contributions

N, SDR, RRA, and MNS conceived the study. N was analysed under the supervision of RRA and MNS. The first drafts were prepared by N with guidance from SDR, RRA, MNS, and all authors contributed to critical revisions and approved the final manuscript.

## Declarations

### Ethics Approval and Consent to Participate

This study received ethical clearance from the Research Ethics Committee, Faculty of Medicine, Public Health, and Nursing, Universitas Gadjah Mada, Yogyakarta, Indonesia (KE/FK/0182/EC/2025). Written and verbally informed consent was obtained from all participants before data collection, and the study was conducted in accordance with national and international ethical standards.

### Limitations of the Study

This study is subject to potential information bias regarding the variables of completeness and timeliness, as data collection was conducted several months after children had received PCV immunization. In addition, some respondents were caregivers other than the parents, which may have introduced misclassification bias, particularly for variables related to parental knowledge, family support, healthcare worker support, and acceptance of multiple injections.

### Consent for Publication

This manuscript does not have personal identification; thus, consent for publication was not applicable

## Supporting Information

**S1 Fig. Barriers to PCV Immunization Completeness and Timeliness among Respondents**. This figure compares the reasons cited by caregivers for incomplete immunization (red bars) versus complete but untimely immunization (blue bars). It illustrates the prevalence of specific barriers—including child illness, lack of awareness, forgetfulness, and logistical issues (such as vaccine stock-outs and facility schedules)—highlighting the distinct challenges associated with each immunization outcome.

**S1 Table. Completeness, timeliness, and acceptance of multiple injections for PCV immunization**. This table presents the detailed distribution of immunization coverage for each specific dose (PCV1, PCV2, and PCV3), including the calculated dropout rates from the first to the third dose. Furthermore, it details the timeliness of receipt for each scheduled dose and provides the frequency distribution of caregiver acceptance regarding multiple injections, along with the statistical association between caregiver attitude and acceptance behaviour.

**S2 Table Bivariate analysis of factors associated with completeness and timeliness of PCV immunization**. This table presents the comprehensive results of the bivariate logistic regression analysis. It details the unadjusted associations (Crude Odds Ratios and 95% Confidence Intervals) between all independent variables—including sociodemographic characteristics, perinatal conditions, parental knowledge, support systems, and attitudes—and the two primary outcomes (immunization of PCV completeness and timeliness).

**S1 Dataset. Anonymized primary data set**. This file contains the minimal data set underlying the findings of this study. The Excel workbook consists of two sheets: the “Dataset” sheet providing the coded responses for all 398 participants, and the “Codebook” sheet detailing the variable definitions and coding schemes used in the analysis.

**S1 Questionnaire. Inclusivity in global research**. This checklist outlines the ethical, cultural, and scientific considerations specific to inclusivity in global research. It details the involvement of local researchers, the ethical approval process, and the engagement with local communities and health authorities in the study design and implementation, in accordance with PLOS’ policy on inclusivity in global research.

**Fig 1.**
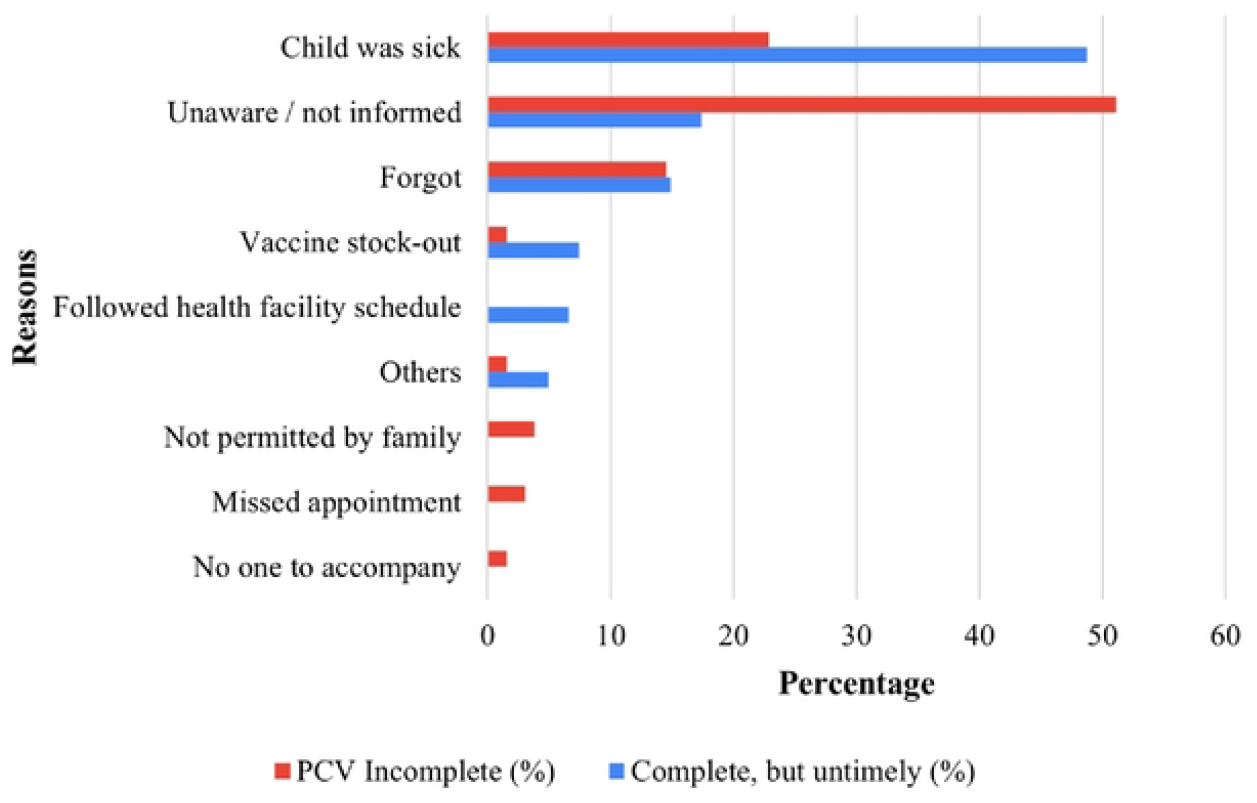
Barriers to PCV Inmmunization Completeness and Timeliness among Respondents.

